# Categorizing ultra-processed food intake in large-scale cohort studies: evidence from the Nurses’ Health Studies, the Health Professionals Follow-up Study, and the Growing Up Today Study

**DOI:** 10.1101/2021.02.08.21251384

**Authors:** Neha Khandpur, Sinara Rossato, Jean-Philippe Drouin-Chartier, Mengxi Du, Euridice Martinez, Laura Sampson, Carlos Monteiro, Fang Fang Zhang, Walter Willett, Teresa T. Fung, Qi Sun

**Author notes:** **Correspondence:** Neha Khandpur, Department of Nutrition, School of Public Health, University of São Paulo, Av. Dr. Arnaldo, 715, São Paulo, Brazil.

## Abstract

**Objective:** There is limited description and documentation of the methods used for the categorization of dietary intake according to the NOVA classification, in large-scale cohort studies. This manuscript details the strategy employed for categorizing the food intake, assessed using food frequency questionnaires (FFQs), of participants in the Nurses’ Health Studies (NHS) I and II, the Health Professionals Follow-up Study (HPFS), and the Growing Up Today Studies (GUTS) I and II into the four NOVA groups to identify the ultra-processed portion of their diets.

**Methods:** A four-stage approach was employed: (1) compilation of all food items from the FFQs used at different waves of data collection; (2) assignment of food items to a NOVA group by three researchers working independently; (3) checking for consensus in categorization and shortlisting food items for which there was disagreement; (4) discussions with experts and use of additional resources (research dieticians, cohort-specific documents, online grocery store scans) to guide the final categorization of the short-listed items.

**Results:** At stage 1, 205 and 315 food items were compiled from the adult and GUTS FFQ food lists, respectively. Over 70% of food items from all cohorts were assigned to a NOVA group after stage 2 and the remainder were shortlisted for further discussion (stage 3). Two rounds of reviews at stage 4 helped with the categorization of 96.5% of items from the adult cohorts and 90.7% items from the youth cohort. The remaining products were assigned to a non-ultra-processed food group and ear-marked for sensitivity analyses. Of all items in the food lists, 36.1% in the adult cohorts and 43.5% in the GUTS cohorts were identified as ultra-processed.

**Conclusion:** An iterative, conservative approach was used to categorize food items from the NHS, HPFS and GUTS FFQ food lists according to their grade of processing. The approach relied on discussions with experts and was informed by insights from the research dieticians, information provided by cohort-specific documents, and scans of online supermarkets. Future work is needed to validate this approach.

## Introduction

Observational epidemiologic studies are invaluable for improving our understanding of the complex relationship that exists between dietary intake and health outcomes. They have helped uncover the role of individual dietary components [1], of specific food groups [2], as well as that of broader dietary patterns [3], in disease etiology and prevention [4]. More recently, observational studies have provided the first evidence for the health consequences associated with the intake of ultra-processed foods including positive associations with the risk of metabolic diseases, cardiovascular disease, depression, and all-cause mortality [5-7]. Ultra-processed foods are one of the four categories of foods that make up the NOVA classification system [8]. They are ready-to-eat/heat industrial formulations of food substances that have been derived from whole foods, and that typically contain added flavours, colours, and other cosmetic additives.

For the most part, the large-scale prospective cohort studies that have assessed the associations between ultra-processed foods and disease outcomes have used self-administered food frequency questionnaire (FFQ) for repeated dietary assessment [9 – 13]. FFQs reflect the mix of food items that constitute the dietary composition and the ultra-processed fraction of the diet, consumed over a designated period of time (usually a year). FFQs are cheaper to administer and easier to comply with, and the usual dietary intake assessed by FFQs often represents a more relevant exposure to chronic disease etiology than dietary intake assessed by methods such as 24-hour diet recalls and records (not usual intake unless assessed repeatedly) [14]. However, the food lists on the basis of which dietary information is collected, have a limited number of pre-defined items which represent the primary sources of energy and nutrients in the population under study. As such, FFQs are unable to cover the full spectrum of ultra-processed foods consumed. Additionally, all supporting information that would be useful in identifying the ultra-processed products from the food lists like cooking methods used, place of food consumption, and brand names of packaged products are usually not captured by FFQs.

It is also worth noting that capturing ultra-processed food intake may not have been the explicit goal of epidemiologic studies at the time of their inception. This would have implications for the development of their dietary assessment instruments. Even 24-hour diet recalls or diet records, that describe with some detail the foods eaten and their method of preparation, may therefore still lack the granularity needed to accurately identify all ultra-processed products. These limitations result in some ambiguity in the identification of the grade of processing of a subset of food items irrespective of the dietary assessment method used, creating an opportunity for a discussion of the possible approaches for improving the identification and estimation of ultra-processed foods in epidemiologic studies.

The purpose of this manuscript is to detail the strategy employed for categorizing the food intake of participants in the Nurses’ Health Studies I and II, the Health Professionals Follow-up Study, and the Growing Up Today Study I and II, collected using semi-quantitative FFQs, into the four NOVA groups to identify the ultra-processed portion of their diets. Collectively, these cohorts have made important contributions to the recommendations of dietary guidelines and nutrition policy [15], and their semi-quantitative FFQs have served as a template for FFQs used in epidemiologic studies across the globe [16-18]. Presenting the approach adopted in these cohorts will inform the categorization of dietary intake in other studies that have used similar assessment methods. While no validation work is presented, the broader goal of this manuscript is to encourage discussions on approaches to categorize dietary data into the NOVA groups and to inform the evolution of dietary assessment methods.

## Methodology

### Cohort details

The Nurses’ Health Study (NHS) began in 1976 with the enrollment of 121,701 female registered nurses who were between 30 and 55 years at the time [19]. In 1986, 51,529 male health professionals between the ages of 40 and 75 years were enrolled into the first cohort of the Health Professionals Follow-Up Study [20], and soon after in 1989, the first cohort of the NHS-II commenced with 116,686 female registered nurses who were between 25 - 42 years. Participants in all three adult cohorts complete a follow-up questionnaire every 2 years (response rate of ∼ 90% at each wave) on their medical history, lifestyle factors, and occurrence of chronic diseases.

In 1996, the children of the NHS-II study participants were recruited into a study of their own – the Growing Up Today Study (GUTS) [21]. At inception, GUTS included 16,882 girls and boys between the ages of 9 and 14 years. In 2004, the study expanded to include a second cohort of 10,920 children between the ages of 10 and 17 - the GUTS-II cohort. The two youth cohorts were surveyed separately and biannually until 2013 when they were merged.

### Assessment of food intake

Dietary data from all waves of both the adult and the youth cohorts were collected using a self-administered, semi-qualitative FFQ [19, 21]. The first 116-item-FFQ was administered to the NHS participants in 1984 to obtain information on usual intake of food and beverages. The FFQ was expanded in 1986 to ∼ 130 food items and sent every 4 years thereafter to participants in both the NHS and the HPFS cohorts (since 1991 in the NHS-II). The GUTS FFQ was modified from the validated FFQ used in the adult cohorts to specifically include snack foods consumed by a younger population and food eaten away from home [21]. The ∼ 150 food item-FFQ was tailored to the cognitive level and dietary knowledge of adolescents. The FFQs have continued to be updated to capture more detailed information on cooking methods and relevant food items [19].

All FFQs ask participants how often, on average, they consumed a given reference portion of a food item over the course of the previous year. Nine response categories capture usual intake, ranging from “never or less than once/month” to “≥6 times/day.” Nutrient intakes are computed by assigning a daily frequency weight [22]. The reproducibility and validity of these FFQs has been extensively evaluated [20-23]. A team of research dietitians maintains an updated database of the nutrient content of foods included on all the FFQ, that reflect the changes to nutrient composition over time (e.g., changes in *trans* fat content) by researching the nutrient content of new or reformulated products [19]. The compilation of the Harvard nutrient database began in 1984, and it has been updated every 4 years to include new food items and the most recent information on food components based on the U.S. Department of Agriculture (USDA) Nutrient Database for Standard Reference and the Food and Nutrients Database for Dietary Studies. The latest version of the Harvard Nutrition Service Center’s nutrient database was issued in 2014. It accounts for 692 food items, recipes, and their ingredients with associated information on 230 food components including total calories, macronutrients, alcohol, sterols, amino acids, fatty acids, vitamins, and minerals [24].

### Classification of food intake into the NOVA groups

The NOVA classification considers the extent and purpose of processing of the food item. The category of ultra-processed foods normally undergo industrial processing like hydrolysis, or hydrogenation, extrusion, moulding, and pre-frying. The three other categories that make up the NOVA classification include unprocessed or minimally processed food, processed culinary ingredients, and processed foods. Compared to ultra-processed foods, these NOVA categories include food products that have undergone processing methods like grinding, roasting, pasteurization, freezing, vacuum packaging or non-alcoholic fermentation (minimally processed foods), centrifuging, refining, or extracting (processed culinary ingredients) or preservation methods such as canning and bottling (processed foods) [8].

A four-stage process was undertaken to identify the ultra-processed foods from the food lists in both the adult and the youth FFQs. First, all food items for which information was collected across different waves of data collection were complied. Food items that were nearly identical between FFQs, but were presented with minor differences (e.g., *“Cold breakfast cereal (1 bowl)”* and *“Cold breakfast cereal (1 serving)”*) were captured as separate items. This was done to make sure that no food item was overlooked. FFQs from every 4 years of the NHS-I (1986 – 2010), the NHS-II (1991 – 2015), the HPFS (1986 – 2014), from 1996, 1998, 2001 for GUTS-I and from 2004, 2006, 2008, 2011 for GUTS-II were used.

Second, three researchers working independently assigned foods in the adult (N.K, S.R, E.M) and the youth (N.K, M.D, E.M) cohorts to one of the four NOVA groups based on their grade of processing – unprocessed / minimally processed foods (G1), processed culinary ingredients (G2), processed foods (G3), and ultra-processed foods (G4). Categorization was an iterative process requiring the review of the original FFQs used to gather dietary information at each wave of data collection to contextualize food items within the larger food lists. At the third stage, categorization between researchers was triangulated. Food items for which there was consensus in the categorization among all researchers were assigned to their NOVA group. A food item was flagged for further scrutiny in case of disagreement in categorization by any two researchers.

At stage four, an expert panel, comprising of three senior researchers (F.F.Z; T.F; Q.S) with substantial experience working with the dietary intake in these cohorts, was convened to review and discuss the categorization of the short-listed products. All discussions were additionally informed by the following resources:

1. Consultations with the research dietitians. The team of research dietitians, led by L.S, was responsible for overseeing the collection of dietary data and for ascertaining the nutrient composition of food items across all Harvard cohorts. They shared their insights from gathering supplementary dietary data, keeping track of product launches and product reformulation of items available in the food retail environment, and conducting multiple pilot studies with participants.
2. Cohort-specific documents. These resources provided information on recipes for some food preparations, or further detail on the key aspects of the food items (a more detailed description, brand name of certain packaged foods).
3. Supermarket scans. The ingredient lists of the first five brands of specific products that were displayed on the Walmart website in 2019 and 2020 were scrutinized and served as a proxy for establishing the level of processing for a small proportion of food items for which limited information was available from the resources listed above.

The process of categorization of food items at this stage was also iterative and at the end of stage four, all products were categorized into one of the four NOVA groups. The compilation and categorization of food items from both the adult and the youth cohorts was done in Microsoft Excel (Microsoft 365, academic license).

## Results

At stage one, total of 205 unique foods from all FFQ food lists of the NHS, NHS-II, and HPFS cohorts and 315 foods in the GUTS cohort were identified and compiled. These included individual food items (“*Butter”; “Coffee”; “Prunes”; “White rice”*) and food items that were combined into a food category. Ninety seven percent of the food items in the adult cohorts (n=199 of 205) and 93% in the GUTS cohort (n= 293 of 315) were asked in this manner. Of the food items in a food category, a large majority of them had similar grades of processing. Examples include “*Tangerines, clementines, mandarin oranges”, “English muffins, bagels, rolls”, “Shrimp, lobster, or scallops as a main dish”, “Beef, pork hotdog”*. Some food categories included specific examples like *“Cereal/Granola bar like Nature Valley, Quaker, or Special K”, “Hot breakfast cereal, like oatmeal, grits”, “Non-fat iced coffee dairy drinks, like Coffee Coolatta, Frappuccino”*. However, about 3% (n=6 of 205) of the food items in the NHS and the HPFS cohorts and 7% (n=22 of 315) in the youth cohort, included a combination of food items with potentially different grades of processing. For instance, in *“Jams, jellies, preserves, honey”*, honey would be differently processed from the other condiments. Similarly, for *“Pie, home-baked or ready-made”*, home-made pies would be differently processed to ready-made pies. Other examples of food categories which grouped food items with different grades of processing include *“Onion rings, cooked onions, or soup”, “Tofu, soyburgers, miso, edamame, or other soy dish”*.

Food items were assigned to a NOVA group by three researchers in stage two. At this stage, cohort- and year-specific FFQs were also used to inform categorization. For instance, the classification of “*Cold breakfast cereal”* into G4, was informed by contextualizing it relative to another item on the FFQ, “*Cooked oatmeal, oatbran”* (G1), for which information was also collected in the same year.

NOVA group assignments were triangulated in stage 3. There was consensus among all study researchers in the categorization of 144 of the 205 food items (70.2%) in the adult cohorts and 221 of the 315 food items (70.2%) in the youth cohort. For example: “*Rice”, “Celery”, “Raw carrot”* were assigned to G1; *“Butter”, “Canola oil”* to G2; *“Canned tuna”, “Olives”* to G3; and *“Jello”, “Ready-made soup from a can”* to G4. There was some discordance in the categorization of 61 of the 205 food items in the adult cohorts (29.7%) and 94 of 315 food items in the youth cohort (29.8%). These included all the food categories with a combination of food items with potentially different grades of processing, mentioned above.

The food items with discordant categorization (n=61 in NHS/HPFS; n=94 in GUTS) were further reviewed by the expert panel, in stage four. Consultations with the dietetic team followed by supermarket scans and team discussions, informed the categorization of 18 of the 61 food items in the adult cohorts and 43 of the 94 food items in the GUTS cohorts. These were subsequently assigned to a NOVA group. Sixteen of the 18 foods in the NHS and HPFS cohorts and 25 of the 43 in the GUTS cohort were identified as having a low potential for contributing to the ultra-processed proportion of the diet – in all possible scenarios of categorization, these products would have been assigned to either the minimally processed or the processed groups. Examples include: *“Apricots”, “Prunes”, “Walnuts”* (G1); *“Sauerkraut”, “Cottage or ricotta cheese”, “Mustard”* (G3). The processing category for *“Mustard”* (G3) was informed by the supermarket scans. Food products (n=2 in NHS/HPFS; n=18 in GUTS) that were classified as ultra-processed at this stage included *“Salad dressing”, “Soy or Worcestershire sauce”, “Salami, bologna, or other deli meat sandwich”, “Tofu, soyburgers, other meat substitutes”, “Hawaiian Punch, lemonade, Koolaid or other non-carbonated fruit drinks”*.

In continued review in stage four, cohort-specific documents, along with further input from the dietetic team, were used to determine the level of processing for 34 additional food items in the adult cohorts and 22 additional food items in the GUTS cohort. These products were subsequently assigned to a NOVA group. For instance, cohort-specific documents described *“Applesauce”* as ‘applesauce, canned, sweetened, and without salt’ and *“Canned peaches”* as ‘peaches, canned, heavy syrup; peaches, canned in juice’ which helped with their assignment into G4. The documents also provided a detailed description for items like *“French fried potatoes”* [frozen french fries prepared, McDonald’s french fries; Burger King french fries]. Additional examples of food items that were assigned to the ultra-processed food group at this stage include: *“Brownies”, “French fried potatoes”, “Pizza”, “Chowder or cream soup”, “Dairy coffee drink”, “Danish, sweet rolls, pastry”*.

There was not enough evidence in the resource documents to support the classification of the 9 remaining food items in the NHS and HPFS cohorts and 29 remaining food items in the GUTS cohort. After discussion with the expert panel, a conservative approach to their categorization was adopted by assigning these products to a non-ultra-processed NOVA group as their primary categorization. These products were also flagged for further sensitivity analysis at which point they would be assigned to the ultra-processed group. Examples of the food items that were categorized in this manner include: *“Popcorn”* (G3); *“Soy milk”* (G1); *“Chicken or turkey sandwich”* (G1); *“Pancakes or waffles”* (G1); *“Pie, home-baked or ready-made”* (G1). A flow chart of the categorization process and cumulative categorized percent is presented in Figure 1.

**Figure 1.**
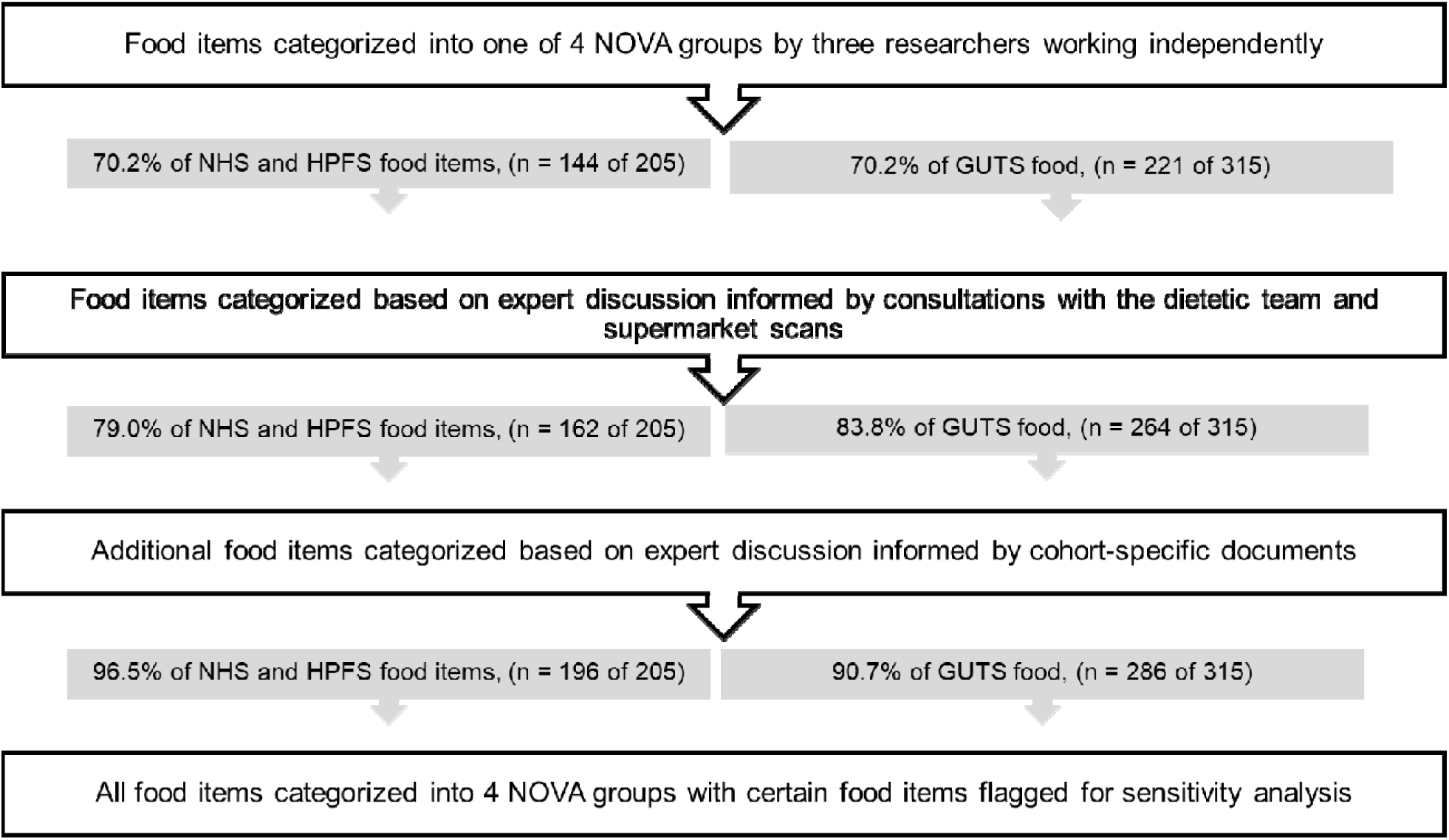
The process of NOVA categorization of dietary data in the Nurses’ Health Studies I and II (NHS), the Health Professionals Follow-up Study (HPFS) and the Growing Up Today Study (GUTS)

A total of 74 of the 205 food items (36.1%) from the NHS and HPFS FFQ food lists and 137 of the 315 food items (43.5%) from the GUTS FFQ food lists were assigned to the ultra-processed food category, at the end of the categorization process at stage 4. Of these, 85.1% of the food item in the adult cohorts (63 of 74) and 72.9% in the youth cohort (100 of 137) were categorized at the end of stage 2, even before discussion with experts (e.g., “*Regular carbonated beverage with caffeine & sugar”*, “*White bread, pita bread, or toast”*, “*Popsicles”*). Over 86% of the ultra-processed foods from all cohorts were categorized after the first round of reviews with experts. Figure 2 presents the contribution of the 4 NOVA groups to the FFQ food lists compiled from all waves of the cohorts.

**Figure 2.**
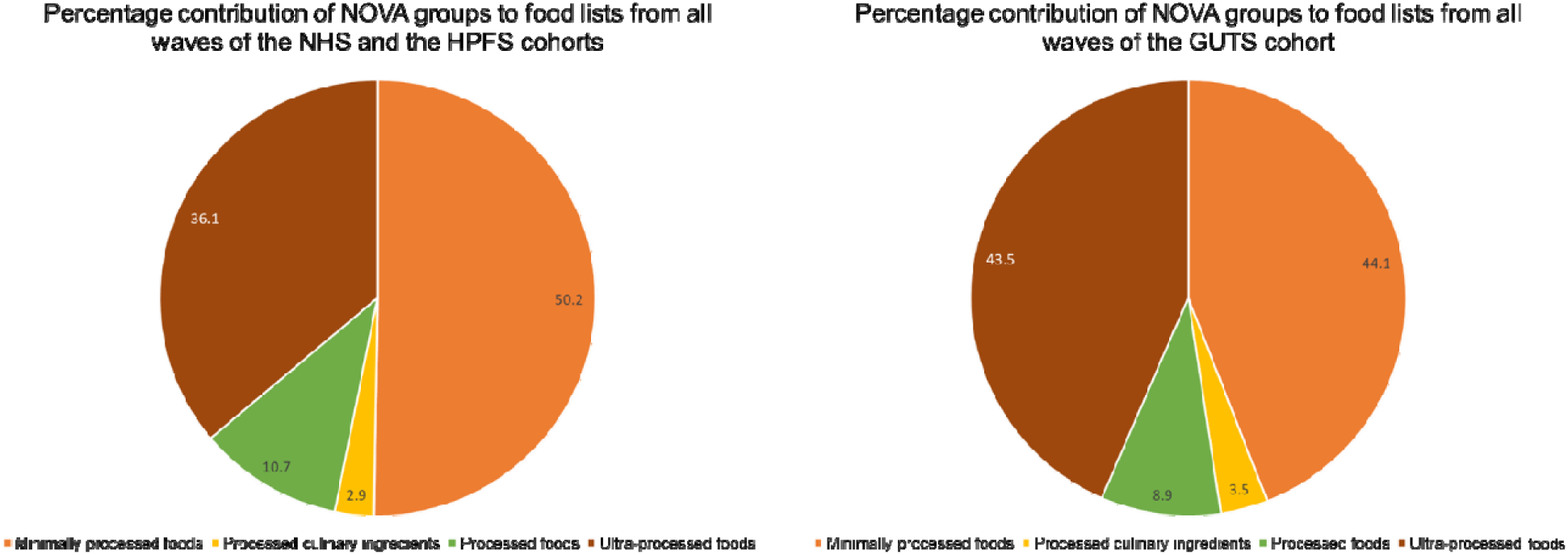
The percentage contribution of the 4 NOVA groups to the food items compiled from all waves of the Nurses’ Health Studies I and II (NHS), the Health Professionals Follow-up Study (HPFS) and the Growing Up Today Study (GUTS)

Table 1 and Table 2 capture the food items scrutinized at stage four.

**Table 1.**
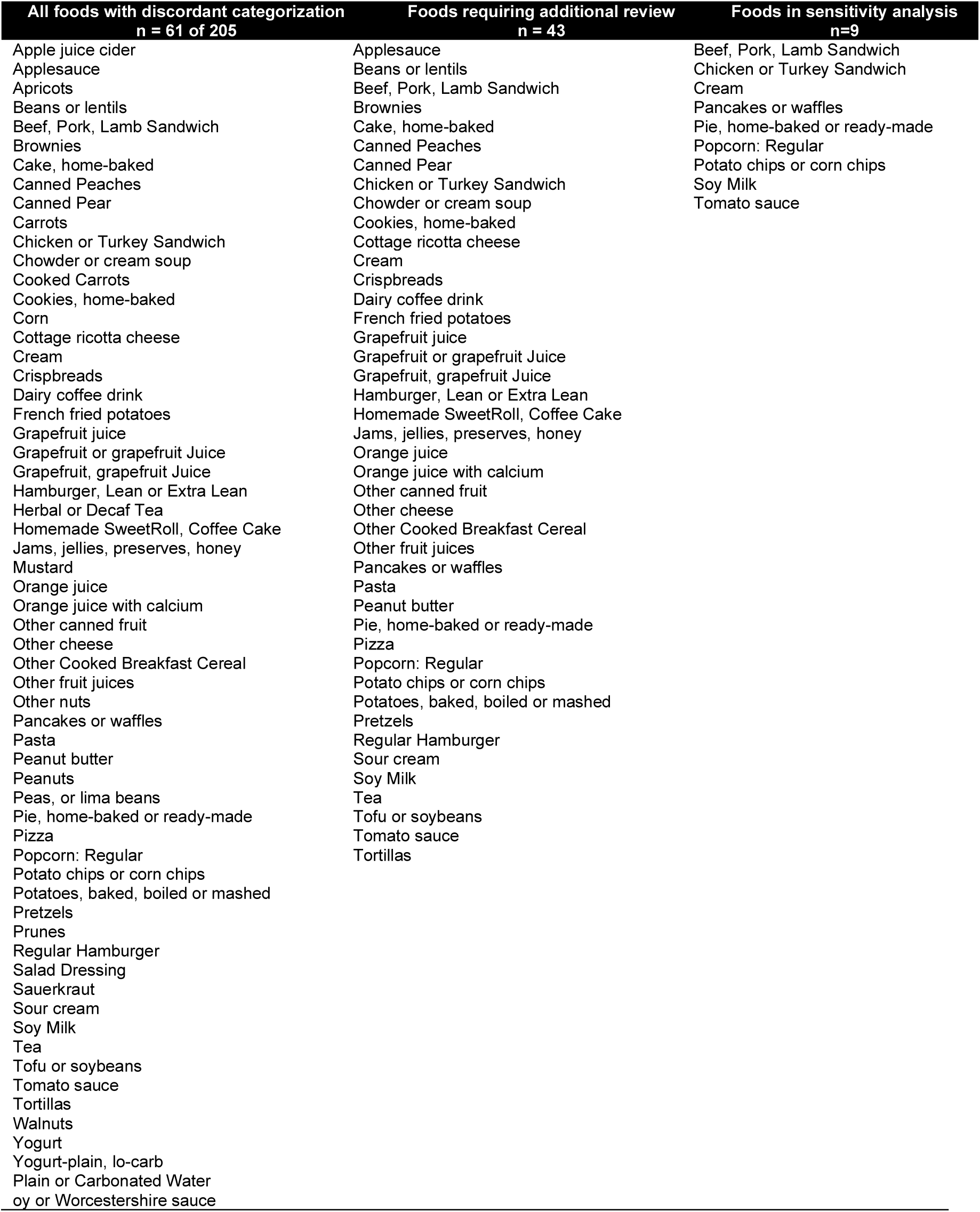
Foods items with discordant categorization that required discussion and further review in the Nurses’ Health Studies I and II (NHS), the Health Professionals Follow-up Study (HPFS)

| All foods with discordant categorization<br>n = 61 of 205 | Foods requiring additional review<br>n = 43 | Foods in sensitivity analysis<br>n=9 |
| --- | --- | --- |
| Apple juice cider | Applesauce | Beef, Pork, Lamb Sandwich |
| Applesauce | Beans or lentils | Chicken or Turkey Sandwich |
| Apricots | Beef, Pork, Lamb Sandwich | Cream |
| Beans or lentils | Brownies | Pancakes or waffles |
| Beef, Pork, Lamb Sandwich | Cake, home-baked | Pie, home-baked or ready-made |
| Brownies | Canned Peaches | Popcorn: Regular |
| Cake, home-baked | Canned Pear | Potato chips or corn chips |
| Canned Peaches | Chicken or Turkey Sandwich | Soy Milk |
| Canned Pear | Chowder or cream soup | Tomato sauce |
| Carrots | Cookies, home-baked |  |
| Chicken or Turkey Sandwich | Cottage ricotta cheese |  |
| Chowder or cream soup | Cream |  |
| Cooked Carrots | Crispbreads |  |
| Cookies, home-baked | Dairy coffee drink |  |
| Corn | French fried potatoes |  |
| Cottage ricotta cheese | Grapefruit juice |  |
| Cream | Grapefruit or grapefruit Juice |  |
| Crispbreads | Grapefruit, grapefruit Juice |  |
| Dairy coffee drink | Hamburger, Lean or Extra Lean |  |
| French fried potatoes | Homemade SweetRoll, Coffee Cake |  |
| Grapefruit juice | Jams, jellies, preserves, honey |  |
| Grapefruit or grapefruit Juice | Orange juice |  |
| Grapefruit, grapefruit Juice | Orange juice with calcium |  |
| Hamburger, Lean or Extra Lean | Other canned fruit |  |
| Herbal or Decaf Tea | Other cheese |  |
| Homemade SweetRoll, Coffee Cake | Other Cooked Breakfast Cereal |  |
| Jams, jellies, preserves, honey | Other fruit juices |  |
| Mustard | Pancakes or waffles |  |
| Orange juice | Pasta |  |
| Orange juice with calcium | Peanut butter |  |
| Other canned fruit | Pie, home-baked or ready-made |  |
| Other cheese | Pizza |  |
| Other Cooked Breakfast Cereal | Popcorn: Regular |  |
| Other fruit juices | Potato chips or corn chips |  |
| Other nuts | Potatoes, baked, boiled or mashed |  |
| Pancakes or waffles | Pretzels |  |
| Pasta | Regular Hamburger |  |
| Peanut butter | Sour cream |  |
| Peanuts | Soy Milk |  |
| Peas, or lima beans | Tea |  |
| Pie, home-baked or ready-made | Tofu or soybeans |  |
| Pizza | Tomato sauce |  |
| Popcorn: Regular | Tortillas |  |
| Potato chips or corn chips |  |  |
| Potatoes, baked, boiled or mashed |  |  |
| Pretzels |  |  |
| Prunes |  |  |
| Regular Hamburger |  |  |
| Salad Dressing |  |  |
| Sauerkraut |  |  |
| Sour cream |  |  |
| Soy Milk |  |  |
| Tea |  |  |
| Tofu or soybeans |  |  |
| Tomato sauce |  |  |
| Tortillas |  |  |
| Walnuts |  |  |
| Yogurt |  |  |
| Yogurt-plain, lo-carb |  |  |
| Plain or Carbonated Water |  |  |
| oy or Worcestershire sauce |  |  |

**Table 2.**
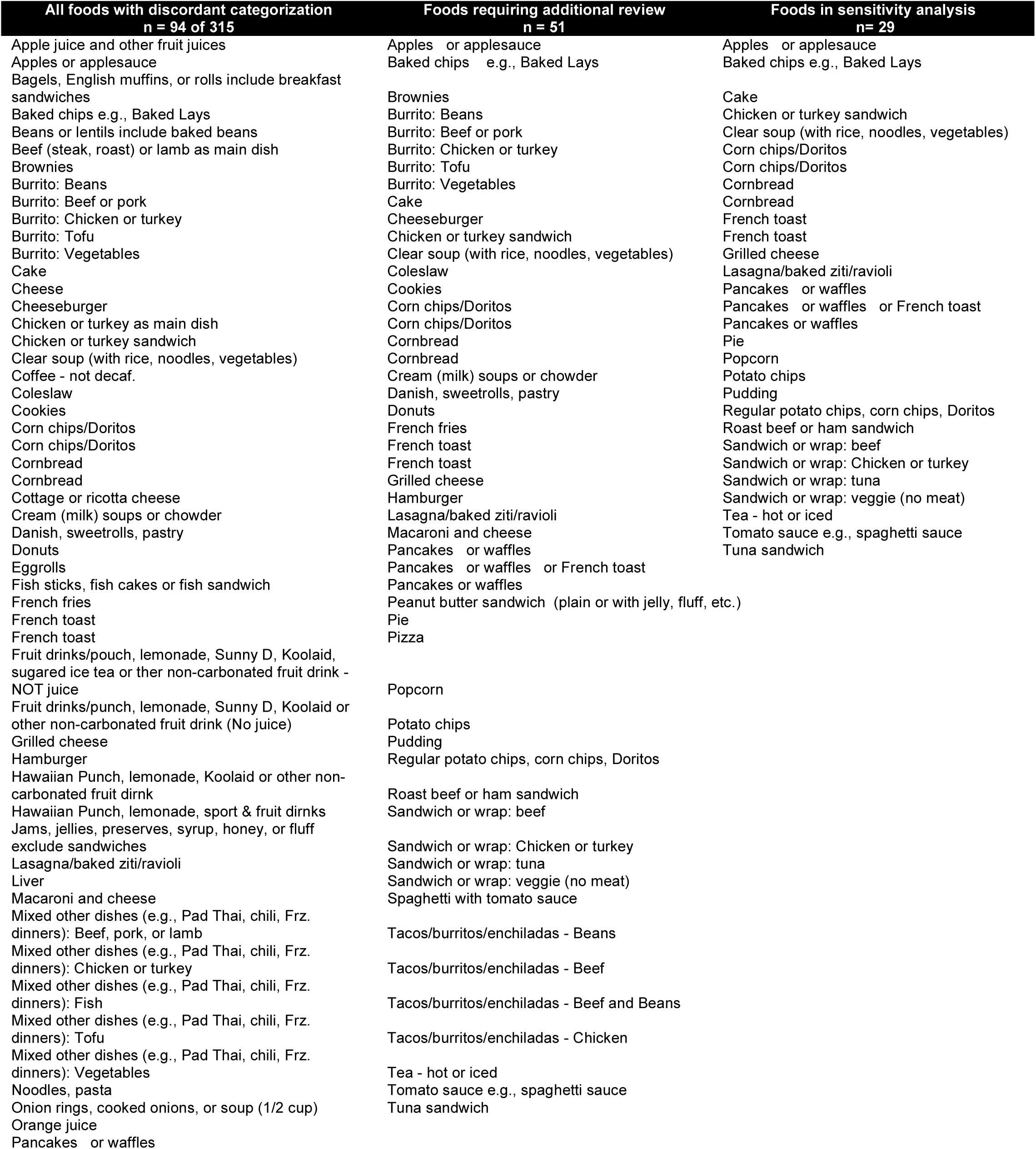
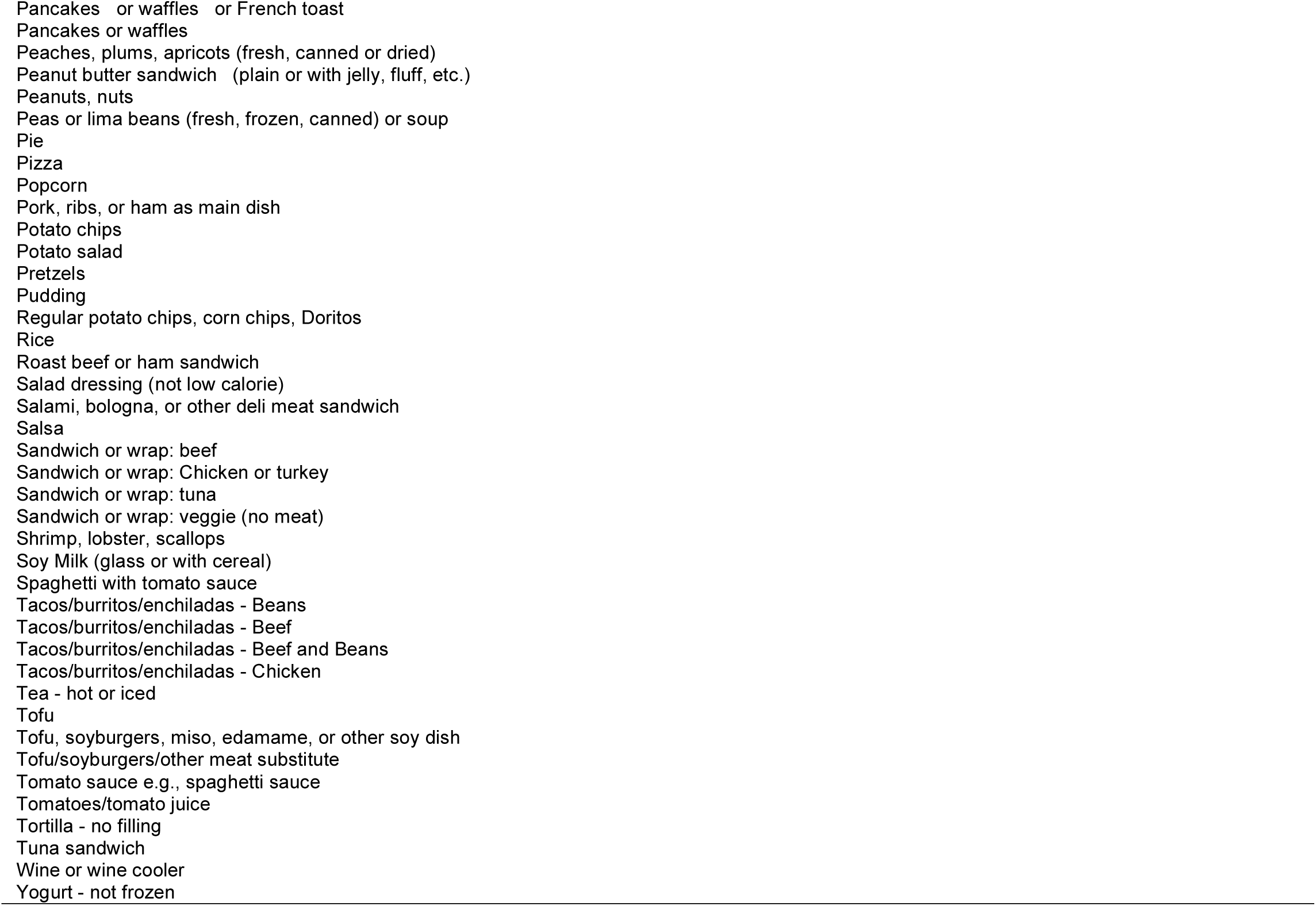
Foods items with discordant categorization that required discussion and further review in the Growing Up Today Study.

The final NOVA classification of food items in the NHS and the HPFS cohorts is presented in Table 3. Table 4 presents the NOVA classification of the GUTS food items. The classification of the dietary intake formed the basis for the development of four different indicators that reflected the portion of ultra-processed foods in the diets – absolute kilocalories from ultra-processed foods, percentage of kilocalories from ultra-processed foods, percentage of grams of ultra-processed foods, and the servings per day from ultra-processed foods.

## Discussion

This article details the process of estimating the ultra-processed portion of the dietary intake data collected using FFQs of participants in the NHS-I and II, the HPFS and the GUTS. Over 70% of the food items across all cohorts were assigned into one of the four NOVA groups after the first attempt at categorization. The approach to classify the remaining 30% of food items involved discussions with experts that were informed by insights from the research dieticians, information provided by cohort-specific documents, and scans of online grocery stores.

A conservative approach to the classification of some of the more challenging food items was adopted, which meant that only food items that could be justifiably considered ultra-processed based on information from cohort-specific documents were assigned to this NOVA category. In the handful of instances of uncertainty, the food items were considered as ultra-processed only for sensitivity analysis and a non-ultra-processed NOVA group was assigned as their primary classification. It is therefore reasonable to assume that the ultra-processed portion of the diet is slightly underestimated and caution needs to be exerted in the interpretation of the absolute intake of UPFs estimated from FFQs. While this may have an attenuating effect on any future diet-disease relationship studied, the variability associated by this approach will be possible to measure via the recategorization planned for sensitivity analyses.

Besides the resources mentioned in this article to help with the NOVA categorization of dietary data, additional resources may be considered. Since there is no one gold-standard for applying the NOVA categorization, potential resources could include the use of year-appropriate, context- or region-specific, nationally representative surveys that use 24-hour diet recalls or diet records to gather more detailed information on dietary intake. The food items from these surveys could help determine the level of processing of certain challenging products. For instance, the National Health and Nutrition Examination Surveys (NHANES) in the US could be used to provide some insight into the processing grade of foods consumed in the adult and youth cohorts if there was no access to cohort-specific resources.

Alternate approaches were considered but not implemented. One option was not taking a conservative approach in assigning certain foods to the ultra-processed category. The other option was to split the food items that could be classified into more than one NOVA group and to use appropriate resources to justify the allocation of a percentage of the food item into each group. For instance, “*Pie, home-baked or ready-made”* could be split to allocate 60% of its nutrient profile to the minimally processed group and 40% to the ultra-processed group. This approach of dividing the nutrient profile of the food item into more than one NOVA group has been done before [10, 25]. It may either be an equal split between different NOVA groups or be divided in a proportion that is informed by other sources [25]. Validation studies will be needed to estimate the misclassification minimized in determining the ultra-processed foods by these approaches. Previous work has examined the performance of the FFQ with diets measured by the average of two 7-day dietary records, and of four web-based automated-self-administered 24-hour recalls kept over a one-year period [26].

Insights from this work could inform the refinement of existing dietary collection instruments to more accurately reflect the level of processing of food items. This may include increasing the specificity of certain food items, including brand names of packaged products where possible, capturing the place of consumption (at restaurant, street-food, take-aways), and the manner of preparation of mixed dishes and the types of ingredients used (from scratch with fresh ingredients, pre-made and frozen using processed ingredients) [27]. For FFQs, it could also mean adding more items or sub-dividing existing ones to differentiate between grades of processing and asking follow-up questions that give a better sense of the overall processing of the dietary pattern [28].

Finally, this work may serve as a protocol for applying the NOVA classification and identifying ultra-processed foods for other large-scale cohort studies. While it presents a specific example of a method for the classification of dietary intake collected using FFQs, the various stages and the decision-points detailed in this manuscript could be modified based on context-specific needs and applied to other studies. This work may also be valuable as a template for authors thinking about documenting and making transparent their method to classification and helping increase the reproducibility of their research.

### Strengths and limitations

The strengths and limitations of this approach are important to highlight. The current approach to classification assumed no changes to the grade of processing of the food item or its food composition over time. The classification of a food product as ultra-processed remained static, irrespective of whether that product was consumed in 1986 (first FFQ cycle in these cohorts) or in 2015 (the latest available FFQ cycle in these cohorts) and the nutrient database from 2014 was used to determine the nutrient composition for all products. Reformulation to the processing and the nutrient profile of the products, therefore, was not captured by the current approach. While it is likely that a small portion of foods switched between NOVA groups over the course of dietary data collection, future work would be needed to capture the evolution of the processing of these products and to estimate their contribution to the ultra-processed portion of the diets. The use of the nutrient database from 2014, likely confers a healthier nutrient composition to products that have been subject to reformulation [29, 30].

One limitation of using supermarket scans is the potential for asynchrony between the collection of the dietary information from participants and the website searches. This may result in the identification of different brands of products from the ones consumed by the participants and/or different levels of processing as gauged from the ingredient lists. In the current approach, ingredient lists of the most popular brands of products from 2019 – 2020 were used to reflect the processing of certain food items consumed from 1986 to 2015. To minimise the potential for misclassification, cohort-specific resources were given priority in informing the NOVA categorization of most food items and the grocery website scans, while helpful, were only used to categorize a handful of products.

The food lists included a few food categories that combined food items from different NOVA groups. While it is likely that the caloric contribution of each item was small, the approach to categorization did not attempt to disaggregate the food category into individual items to estimate the dietary contribution of each of the components. This may have led to some misclassification. Finally, this approach to categorization would not address challenges associated with the linking of food composition databases to the foods captured by different dietary assessment methods. For instance, in the case of mixed dishes and baked goods requiring multiple ingredients, the dietary data are often not disaggregated to reflect the nutrient profile of individual components, some of which may be ultra-processed, but instead reflects the overall nutrient composition of the dish in its entirety. Neither does it address inherent systematic biases associated with the reporting of certain foods, that could impede the accurate estimation of the ultra-processed fraction of the diet. These aspects were beyond the scope of the categorization strategy adopted.

The strengths of this approach were the triangulation of NOVA group assignment, expert review of the categorization process, the use of supporting documents to inform the categorization of the more challenging food items, and the high transferability of this approach to categorizing dietary data collected using other diet assessment methods.

## Conclusions

This manuscript presents the strategy used in the estimation of the ultra-processed portion of the diet of participants in large-scale population studies. The iterative, conservative approach adopted, relied on discussions with experts and was informed by insights from the research dieticians, information provided by cohort-specific documents, and scans of online grocery stores. All food items were assigned a primary NOVA group with some foods being ear-marked for further sensitivity analysis. Future work would be needed to certify the validity of this approach by comparing the UPF intakes estimated through FFQs against diet records and biomarkers. An evaluation of the generalizability and feasibility of applying this approach to other study populations and contexts is also warranted. Documentation of and discussions on alternative approaches for categorization along with their validation, and on the evolution of dietary assessment methods to better capture ultra-processed foods are encouraged.

## Data Availability

All data reported is available on request

## Notes

### Competing Interest Statement

The authors have declared no competing interest.

### Funding Statement

No external funding was received.

### Author Declarations

This manuscript contains no participant data and was considered IRB exempt.

