## Supplementary material for "Categorizing ultra-processed food intake in large-scale cohort studies: evidence from the Nurses’ Health Studies, the Health Professionals Follow-up Study, and the Growing Up Today Study": Table 3

**Table 3. The classification of all food items captured by the semi quantitative food frequency questionnaires used in the Nurses’ Health Studies I and II (NHS), the Health Professionals Follow-up Study (HPFS), into the NOVA groups of minimally processed foods (1), processed culinary ingredients (2), processed foods (3), and ultra-processed foods (4)**

| **Food item** | **NOVA group assignment** |
| --- | --- |
| 1% -2% Milk | 1 |
| Alfafa sprouts | 1 |
| Apple juice cider | 3 |
| Applesauce | 4 |
| Apricots | 1 |
| Avocado | 1 |
| Bacon | 4 |
| Banana | 1 |
| Beans or lentils | 1 |
| Beef, calf, pork liver | 1 |
| Beef, lamb as a main dish | 1 |
| Beef, pork hot dogs | 4 |
| *Beef, pork, lamb sandwich** | 1* |
| Beer | 3 |
| Beets | 1 |
| Blueberries | 1 |
| Breaded fish cakes, pieces, sticks | 4 |
| Breakfast bar | 4 |
| Broccoli | 1 |
| Brown rice | 1 |
| Brownies | 4 |
| Brussels sprouts | 1 |
| Butter | 2 |
| Caffeine free Coke Pepsi | 4 |
| Cake, home-baked | 1 |
| Cake, ready-made | 4 |
| Candy bar with chocolate | 4 |
| Candy bar without chocolate | 4 |
| Canned peaches | 4 |
| Canned pear | 4 |
| Canned tuna | 3 |
| Cantaloupe | 1 |
| Carrots | 1 |
| Cauliflower | 1 |
| Celery | 1 |
| Chicken or turkey hot dogs | 4 |
| Chicken or turkey liver | 1 |
| *Chicken or turkey sandwich** | 1* |
| Chicken or turkey, with skin | 1 |
| Chicken or turkey, without skin | 1 |
| Chocolate bars | 4 |
| Chowder or cream soup | 4 |
| Coffee | 1 |
| Coke, Pepsi cola | 4 |
| Cold breakfast cereal | 4 |
| Cooked cabbage | 1 |
| Cooked carrots | 1 |
| Cooked oatmeal, oatbran | 1 |
| Cookies, brownie ready-made | 4 |
| Cookies, fat free, reduced fat | 4 |
| Cookies, home-baked | 1 |
| Corn | 1 |
| Cottage ricotta cheese | 3 |
| *Cream** | 2* |
| Cream cheese | 4 |
| Crispbreads | 3 |
| Cucumber | 1 |
| Dairy coffee drink | 4 |
| Dark chocolate bars | 4 |
| Dark meat fish | 1 |
| Decaffeinated coffee | 1 |
| Diet nutrition drinks, Slimfast | 4 |
| Doughnuts | 4 |
| Dried cranberries | 1 |
| Eggplant, zucchini, summer squash | 1 |
| Energy bar | 4 |
| Energy or high protein bars | 4 |
| English muffins, bagels, rolls | 4 |
| Ensure, boost or other meal replacement drinks | 4 |
| Fat free popcorn | 4 |
| Fat free, light crackers | 4 |
| Figs | 1 |
| Flavored yogurt without Nutrasweet | 4 |
| French fried potatoes | 4 |
| Fresh apples, pears | 1 |
| Fresh pears | 1 |
| Frozen yogurt, sherbet, icecream | 4 |
| Garlic | 1 |
| Grapefruit | 1 |
| Grapefruit juice | 1 |
| Grapefruit or grapefruit juice | 1 |
| Green, red peppers | 1 |
| Hamburger, lean or extra lean | 1 |
| Hawaiian punch | 4 |
| Herbal or decaf tea | 1 |
| High protein, lowcarb candy bar | 4 |
| Home-made soup with bouillon | 4 |
| Home-made soup without bouillon cube | 1 |
| Homemade sweetroll, coffee cake | 1 |
| Hummus | 3 |
| Ice cream | 4 |
| Iceberg or head lettuce | 1 |
| Jams, jellies, preserves, honey | 4 |
| Kale, mustard, or chard green | 1 |
| Ketchup or red chili sauce | 4 |
| Light beer | 3 |
| Liquor | 4 |
| Low calorie soda, caffeine free | 4 |
| Low calorie soda, Pepsi, 7-up | 4 |
| Low fat, fat free mayonnaise | 4 |
| Margarine | 4 |
| Mixed dried fruit | 1 |
| Mixed dried fruits | 1 |
| Mixed vegetables | 1 |
| Muffins or biscuits | 4 |
| Mushrooms | 1 |
| Mustard | 3 |
| Non-dairy whitener | 4 |
| Nutrasweet or Equal | 4 |
| Oat bran | 1 |
| Oil and vinegar dressing | 2 |
| Olive oil added to food | 2 |
| Olives | 3 |
| Omega-3 fortified including yolk | 1 |
| Onion as a garnish | 1 |
| Onion as a vegetable | 1 |
| Orange juice | 1 |
| Orange juice with calcium | 1 |
| Orange squash | 1 |
| Oranges | 1 |
| Other artificial sweetener | 4 |
| Other bran | 1 |
| Other canned fruit | 3 |
| Other carbon beverage | 4 |
| Other cheese | 3 |
| Other cooked breakfast cereal | 1 |
| Other fish | 1 |
| Other fruit juices | 1 |
| Other grains | 1 |
| Other low calorie carb | 4 |
| Other low calorie cola with caffeine | 4 |
| Other nuts | 1 |
| *Pancakes or waffles** | 1* |
| Pasta | 1 |
| Peach, apricots, plums | 1 |
| Peanut butter | 3 |
| Peanuts | 1 |
| Peas, or lima beans | 1 |
| Pepper | 1 |
| Pie, home-baked | 1 |
| *Pie, home-baked or ready-made** | 1* |
| Pie, ready-made | 4 |
| Pizza | 4 |
| Plain or carbonated water | 1 |
| *Popcorn: regular** | 3* |
| Pork as a main dish | 1 |
| *Potato chips or corn chips** | 3* |
| Potatoes, baked, boiled or mashed | 1 |
| Pretzels | 3 |
| Processed meats, sausage | 4 |
| Prune juice | 1 |
| Prunes | 1 |
| Raisins or grapes | 1 |
| Raw carrots | 1 |
| Ready-made soup from can | 4 |
| Ready-made sweetroll, coffeecake | 4 |
| Red chili sauce | 4 |
| Red wine | 3 |
| Regular crackers | 4 |
| Regular hamburger | 1 |
| Regular mayonnaise | 4 |
| Romaine or leaf lettuce | 1 |
| Rye, pumpernickel bread | 4 |
| Salad dressing | 4 |
| Salami, bologna, processed meat sandwiches | 4 |
| Salsa, picante, taco sauce | 4 |
| Salt | 2 |
| Sauerkraut | 3 |
| Shrimp lobster scallop | 1 |
| Skim or low-fat milk | 1 |
| Snack bars | 4 |
| Sour cream | 2 |
| *Soy milk** | 1* |
| Soy or Worcestershire sauce | 4 |
| Spinach, cooked | 1 |
| Spinach, raw | 1 |
| Splena | 4 |
| Spreadable butter | 4 |
| Strawberries | 1 |
| String beans | 1 |
| Sweet roll fat free or reduced fat | 4 |
| Tangerines, clementines, mandarin oranges | 1 |
| Tea | 1 |
| Toasted breads, bagel or English muffin | 4 |
| Tofu or soybeans | 3 |
| Tomato juice | 3 |
| *Tomato sauce** | 3* |
| Tomato soup | 4 |
| Tomatoes | 1 |
| Tortillas | 3 |
| Uncooked cabbage, coleslaw | 1 |
| Walnuts | 1 |
| Watermelon | 1 |
| Wheat germ | 1 |
| White bread | 4 |
| White rice | 1 |
| White wine | 3 |
| Whole eggs | 1 |
| Whole milk | 1 |
| Whole wheat etc whole grain bread | 4 |
| Yams or sweet potatoes | 1 |
| Yogurt | 1 |
| Yogurt artificially sweetened | 4 |
| Yogurt flavored without nutrasweet | 4 |
| Yogurt-plain, lo-carb | 1 |

** indicates foods to be categorized as ultra-processed (4) for sensitivity analysis*
