## Supplementary material for "Categorizing ultra-processed food intake in large-scale cohort studies: evidence from the Nurses’ Health Studies, the Health Professionals Follow-up Study, and the Growing Up Today Study": Table 4

**Table 4. The classification of all food items captured by the semi quantitative food frequency questionnaire used in the Growing Up Today Study, into the NOVA groups of minimally processed foods (1), processed culinary ingredients (2), processed foods (3), and ultra-processed foods (4)**

| **Food item** | **NOVA group assignment** |
| --- | --- |
| 1 or 2 % milk | 1 |
| 1% milk | 1 |
| 2% milk | 1 |
| Added sugar | 2 |
| Apple juice, other 100% fruit juices | 1 |
| Apple juice and other 100% fruit juices | 1 |
| Apple juice and other fruit juices | 1 |
| *Apples or applesauce* | 1* |
| Apples or pears | 1 |
| Applesauce | 4 |
| Bacon | 4 |
| Bacon | 4 |
| Bagels, English muffins, or rolls include breakfast sandwiches | 4 |
| *Baked chips e.g., Baked Lays* | 3* |
| Bananas | 1 |
| Beans or lentils including baked beans | 1 |
| Beans/lentils/edamame | 1 |
| Beans/lentils/soybeans | 1 |
| Beef (steak, roast) or lamb as main dish | 1 |
| Beef or lamb as a main dish, e.g., as a steak or roast | 1 |
| Beef or pork hot dogs | 4 |
| Beef, pork, or lamb fat | 1 |
| Beef/pork sausage | 4 |
| Beer | 3 |
| Beer, regular | 3 |
| Beets (not greens) | 1 |
| Biscuit | 4 |
| Biscuit/roll | 4 |
| Breakfast bars, e.g., Nutrigrain, granola, Kashi | 4 |
| Broccoli | 1 |
| Broccoli | 1 |
| Brown gravy | 4 |
| Brown rice | 1 |
| Brownies | 4 |
| Brussels sprouts | 1 |
| Burrito: Beans | 4 |
| Burrito: Beef or pork | 4 |
| Burrito: Chicken or turkey | 4 |
| Burrito: Tofu | 4 |
| Burrito: Vegetables | 4 |
| Butter | 2 |
| Butter - not margarine | 2 |
| *Cake* | 1* |
| Cake or snack cakes, e.g., Twinkies | 4 |
| Canned tuna | 3 |
| Canola oil | 2 |
| Cantaloupe | 1 |
| Cantaloupe, melons | 1 |
| Carbonated, low-cal “diet” beverage with caffeine, (e.g., Diet Coke) | 4 |
| Carrots, cooked | 1 |
| Carrots, cooked | 1 |
| Carrots, raw | 1 |
| Carrots, raw | 1 |
| Celery | 1 |
| Celery | 1 |
| Cereal/Granola bar, like Nature Valley, Quaker, or Special K | 4 |
| Cheese | 3 |
| Cheese, eaten alone or added to main dish, sandwich, or quesadilla | 3 |
| Cheeseburger | 4 |
| Chicken nuggets | 4 |
| Chicken or turkey as a main dish (e.g., fried or roasted) with skin, including grounded | 1 |
| Chicken or turkey as a main dish, without skin | 1 |
| Chicken or turkey as main dish | 1 |
| Chicken or turkey hot dogs or sausages | 4 |
| *Chicken or turkey sandwich* | 1* |
| Chicken or turkey skin | 1 |
| Chocolate like Hershey's or M & M's | 4 |
| Chocolate candy like Hershey's, Snickers or M & M's | 4 |
| Chocolate candy like Snickers or M & M's | 4 |
| Chocolate milk | 4 |
| Chocolate milk, or any flavored milk | 4 |
| Chocolate or other flavored milk | 4 |
| Chocolate, e.g., Hershey’s or M&M’s | 4 |
| *Clear soup (with rice, noodles, vegetables)* | 3* |
| Coffee - not decaf. | 1 |
| Coffee drinks - latte, Coolers, Coolatas, Frappuccinos, Mochachinos | 4 |
| Cold breakfast cereal | 4 |
| Cold breakfast cereal | 4 |
| Coleslaw | 4 |
| Collard greens/kale | 1 |
| Collard greens/kale/cooked spinach | 1 |
| Cooked oatmeal, including instant | 1 |
| Cookies | 4 |
| Corn | 1 |
| Corn | 1 |
| *Corn chips/Doritos* | 3* |
| Corn oil | 2 |
| *Cornbread* | 3* |
| Cornbread | 3 |
| Cottage cheese | 3 |
| Cottage or ricotta cheese | 3 |
| Crackers, e.g., Cheez-its or Ritz | 4 |
| Crackers, like Wheat Thins or Ritz | 4 |
| Cream (milk) soups or chowder | 1 |
| Cream cheese | 4 |
| Cream cheese | 4 |
| Cream, e.g., coffee, sour (exclude fat free) | 2 |
| Danish, donut, sweetroll or pastry | 4 |
| Danish, sweetrolls, pastry | 4 |
| Dark bread | 4 |
| Dark meat fish, e.g., tuna steak, salmon, sardines, swordfish | 1 |
| Decaffeinated coffee | 1 |
| Diet soda | 4 |
| Donuts | 4 |
| Eggrolls | 1 |
| Eggs | 1 |
| Eggs e.g., scrambled, fried, in breakfast sandwich | 1 |
| Energy bar (like Power or Cliff Bar) | 4 |
| Energy bar, e.g., Powerbar, Clif Bar, Lunabar | 4 |
| Energy drink - (e.g Red Bull), Regular energy drinks | 4 |
| Energy drink - (e.g Red Bull), Sugar free, low calorie, or low carb energy drinks | 4 |
| Energy drink - Red Bull, Rock Star | 4 |
| English muffins or bagels | 4 |
| Fish sticks, fish cakes or fish sandwich | 4 |
| French fries | 4 |
| French fries | 4 |
| *French toast* | 1* |
| Fresh fish as main dish | 1 |
| Frozen yogurt | 4 |
| Frozen yogurt or low-fat ice cream | 4 |
| Fruit drinks/pouch, lemonade, Sunny D, Koolaid, sugared ice tea or other non-carbonated fruit drink - NOT juice | 4 |
| Fruit snacks or fruit rollups | 4 |
| Fun fruit or fruit rollups | 4 |
| Graham crackers | 4 |
| Grapes | 1 |
| Green/red peppers | 1 |
| Green/red/yellow peppers | 1 |
| Greens/kale | 1 |
| *Grilled cheese* | 3* |
| Hamburger | 4 |
| Hawaiian Punch, lemonade, Koolaid or other non-carbonated fruit drink | 4 |
| Hawaiian Punch, lemonade, sport & fruit drinks | 4 |
| High protein bar (like MetRx or Balance Bar) | 4 |
| High protein bar, e.g., Zone or Balance Bar | 4 |
| High protein shake or drink, e.g., whey or soy | 4 |
| Hot breakfast cereal, like oatmeal, grits | 1 |
| Hot dogs | 4 |
| Hot tea | 1 |
| Ice cream | 4 |
| Ice cream | 4 |
| Iced tea - sweetened | 4 |
| Instant Breakfast | 4 |
| Instant breakfast drink | 4 |
| Instant Breakfast Drink/High Protein Shake or Drink | 4 |
| Jams, jellies, preserves, syrup, honey, or fluff, exclude sandwiches | 4 |
| Jello | 4 |
| Ketchup | 4 |
| Ketchup | 4 |
| *Lasagna/baked ziti/ravioli* | 1* |
| Lettuce/tossed salad | 1 |
| Lettuce/tossed salad | 1 |
| Light beer, e.g., Bud Light or Natural Light | 3 |
| Liquor, like vodka or rum | 4 |
| Liver | 1 |
| Low calorie or low fat salad dressing | 3 |
| Low calorie or low fat salad dressing | 3 |
| Low-fat or whole milk coffee dairy drinks, e.g., Cappuccino, Mocha, Latte | 4 |
| Low-fat or whole milk iced coffee dairy drinks, e.g., Coffee Coolatta, Frappuccino | 4 |
| Macaroni and cheese | 1 |
| Margarine - not butter | 4 |
| Mayonnaise | 4 |
| Mayonnaise | 4 |
| Meatballs or meatloaf | 1 |
| Milk | 1 |
| Milkshake or frappe | 4 |
| Mixed other dishes (e.g., Pad Thai, chili, Frz. dinners): Beef, pork, or lamb | 4 |
| Mixed other dishes (e.g., Pad Thai, chili, Frz. dinners): Chicken or turkey | 4 |
| Mixed other dishes (e.g., Pad Thai, chili, Frz. dinners): Fish | 4 |
| Mixed other dishes (e.g., Pad Thai, chili, Frz. dinners): Tofu | 4 |
| Mixed other dishes (e.g., Pad Thai, chili, Frz. dinners): Vegetables | 4 |
| Mixed vegetables | 1 |
| Mixed vegetables | 1 |
| Muffin | 4 |
| Nachos with cheese | 4 |
| Nonfat coffee dairy drinks, e.g., Cappuccino, Mocha, Latte | 4 |
| Nonfat iced coffee dairy drinks, e.g., Coffee Coolatta, Frappuccino | 4 |
| Noodles, pasta | 1 |
| Oatmeal | 1 |
| Oatmeal and other hot breakfast cereal, like farina, grits | 1 |
| Olive Oil | 2 |
| Onion rings, cooked onions, or soup | 1 |
| Onions as a garnish or in salad | 1 |
| Orange juice | 1 |
| Orange juice | 1 |
| Oranges, grapefruit | 1 |
| Other candy bars (Milky Way, Snickers) | 4 |
| Other candy bars, e.g., Milky Way, Snickers | 4 |
| Other candy without chocolate (Skittles) | 4 |
| Other carbonated, low-cal beverage without caffeine, (e.g., Diet 7-Up) | 4 |
| Other cooked breakfast cereal | 1 |
| Other fish, e.g., cod, haddock, halibut | 1 |
| Other grains, like kasha, couscous, bulgur | 1 |
| Other hot breakfast cereal, like farina or grits | 1 |
| Other nuts | 1 |
| Other regular carbonated beverage with sugar, (e.g., 7-Up) | 4 |
| *Pancakes or waffles* | 1* |
| *Pancakes or waffles or French toast* | 1* |
| Pasta (e.g., spaghetti with sauce, lasagna): Chicken or turkey | 1 |
| Pasta (e.g., spaghetti with sauce, lasagna): Vegetable | 1 |
| Pasta (e.g., spaghetti with sauce, lasagna): Beef/hamburger/pork | 1 |
| Pasta (e.g., spaghetti with sauce, lasagna): Plain | 1 |
| Peaches, plums, apricots | 1 |
| Peaches, plums, apricots (fresh, canned or dried) | 1 |
| Peanut butter sandwich (plain or with jelly, fluff, etc.) | 4 |
| Peanut butter, plain exclude sandwiches | 1 |
| Peanuts | 3 |
| Peanuts, nut | 3 |
| Pears | 1 |
| Peas or lima beans | 1 |
| Peas or lima beans or soup | 1 |
| *Pie* | 1* |
| Pizza | 4 |
| *Popcorn* | 3* |
| Popsicles | 4 |
| Poptarts | 4 |
| Pork, ribs, or ham as main dish | 1 |
| Posicles | 4 |
| *Potato chips* | 3* |
| Potato salad | 1 |
| Potato salad | 1 |
| Potatoes - baked, boiled, mashed | 1 |
| Potatoes—baked or boiled, mashed | 1 |
| Pretzels | 3 |
| *Pudding* | 1* |
| Pudding - EXCLUDE sugar free | 4 |
| Raisins | 1 |
| Regular carbonated beverage with caffeine & sugar, (e.g., Coke, Pepsi, Mt. Dew, Dr. Pepper) | 4 |
| Regular hot tea with caffeine, including green tea | 1 |
| *Regular potato chips, corn chips, Doritos* | 3* |
| Rice | 1 |
| *Roast beef or ham sandwich* | 1* |
| Safflower oil | 2 |
| Salad dressing (not low calorie) | 1 |
| Salad dressing (not low calorie) | 1 |
| Salami, bologna, or other deli meat sandwich | 4 |
| Salsa | 1 |
| Salsa | 1 |
| *Sandwich or wrap: beef* | 1* |
| *Sandwich or wrap: Chicken or turkey* | 1* |
| Sandwich or wrap: Peanut butter & jelly/fluff | 4 |
| Sandwich or wrap: Salami, bologna, or other deli meat | 4 |
| *Sandwich or wrap: tuna* | 1* |
| *Sandwich or wrap: veggie (no meat)* | 1* |
| Sausage | 4 |
| Seeds (Sunflower or Pumpkin) | 1 |
| Seeds, e.g., sunflower or pumpkin | 1 |
| Shrimp, lobster or scallops as a main dish | 1 |
| Shrimp, lobster, scallops | 1 |
| Skim/nonfat milk | 1 |
| Skim/nonfat milk (glass or with cereak) | 1 |
| Smoothies (e.g., medium Jamba Juice or Orange Julius) | 1 |
| Snack cakes, like Ring Dings/Swiss Rolls/Twinkies | 4 |
| Snack cakes, like Twinkies | 4 |
| Soda - not diet | 4 |
| Soda - not diet | 4 |
| Soy milk (glass or with cereal) | 4 |
| Soy milk, any flavor | 4 |
| Soybean oil | 2 |
| Spaghetti or other pasta with tomato sauce | 1 |
| Spaghetti with tomato sauce | 1 |
| Spinach | 1 |
| Spinach, raw as in salad | 1 |
| Sport drinks - Powerade or Gatorade | 4 |
| Spreadable butter (butter mixed with oil to make it soft and spreadable) | 4 |
| Strawberries | 1 |
| String beans | 1 |
| String beans | 1 |
| Tacos/burritos/enchiladas | 4 |
| Tacos/burritos/enchiladas - Beans | 4 |
| Tacos/burritos/enchiladas - Beef | 4 |
| Tacos/burritos/enchiladas - Beef and Beans | 4 |
| Tacos/burritos/enchiladas - Chicken | 4 |
| *Tea - hot or iced* | 1* |
| Tofu | 3 |
| Tofu, soyburgers, miso, edamame, or other soy dish | 4 |
| Tofu/soyburgers/other meat substitute | 4 |
| Tomato juice or V8 | 1 |
| *Tomato sauce e.g., spaghetti sauce* | 3* |
| Tomatoes | 1 |
| Tomatoes | 1 |
| Tomatoes/tomato juice | 1 |
| Tortilla - no filling | 3 |
| Tortilla e.g., tacos, quesadillas (exclude burritos) | 3 |
| *Tuna sandwich* | 3* |
| Vegetable oil | 2 |
| Veggieburger | 4 |
| Walnuts | 1 |
| Water - tapped or bottled | 1 |
| Watermelon | 1 |
| Wheat or Dark bread | 4 |
| Whipped cream | 4 |
| Whipped cream - EXCLUDE coffee drinks and/or fat free | 2 |
| White bread, pita bread, or toast | 4 |
| White or pita bread, exclude sandwiches | 4 |
| White Rice | 1 |
| Whole milk | 1 |
| Whole milk (glass or with cereal) | 1 |
| Whole wheat or whole grain bread, include toast | 4 |
| Whole wheat or whole grain, exclude sandwiches | 4 |
| Wine or wine cooler | 4 |
| Yams/sweet potatoes | 1 |
| Yams/sweet potatoes | 1 |
| Yogurt - not frozen | 4 |
| Yogurt - plain, not frozen | 1 |
| Yogurt - artificially sweetened (e.g, light peach) | 4 |
| Yogurt - sweetened (e.g., strawberry, vanilla) | 4 |
| Zucchini, summer squash, eggplant | 1 |

** indicates foods to be categorized as ultra-processed (4) for sensitivity analysis*

*Items will not add up to 315 – only one of two or more near identical items [e.g Corn chips/Doritos; Corn chips or Doritos] have been presented here*
